# Physician experiences of electronic health records interoperability and its practical impact on care delivery in the English NHS: A cross-sectional survey study

**DOI:** 10.1101/2024.07.25.24311018

**Authors:** Edmond Li, Olivia Lounsbury, Mujtaba Hasnain, Hutan Ashrafian, Ara Darzi, Ana Luisa Neves, Jonathan Clarke

## Abstract

**Background:** The lack of interoperability has been a well-recognised limitation associated with the use of electronic health records (EHR). However, less is known about how it manifests for frontline NHS staff when delivering care, how it impacts patient care, and what are its implications on care efficiency.

**Objectives:** (1) To capture the perceptions of physicians regarding the current state of EHRs interoperability, (2) to investigate how poor interoperability affects patient care and safety and (3) to determine the effects on care efficiency in the NHS.

**Methods:** An online survey was conducted to explore how physicians perceived the routine use of EHRs, its effects on patient safety, and impact to care efficiency in NHS healthcare facilities. Descriptive statistics was used to report any notable findings observed.

**Results:** A total of 636 NHS physicians participated. Participants reported that EHR interoperability is rudimentary across much of the NHS, with limited ability to read but not edit data from within their organisation. Negative perceptions were most pronounced amongst specialties in secondary care settings and those with less than one year of EHR experience or lower self-reported EHR skills. Limited interoperability prolonged hospital stays, lengthened consultation times, and frequently necessitated repeat investigations to be performed. Limited EHR interoperability impaired physician access to clinical data, hampered communication between providers, and was perceived to threatened patient safety.

**Conclusion:** As healthcare data continues to increase in complexity and volume, EHR interoperability must evolve to accommodate these growing changes and ensure the continued delivery of safe care. The experiences of physicians provide valuable insight into the practical challenges limited interoperability poses and can contribute to future policy solutions to better integrate EHRs in the clinical environment.

**Public Interest/Lay Summary:** Limited interoperability between EHR systems has been a longstanding problem since the technology’s introduction in NHS England. However, little research has been done to understand the extent of this problem from the perspective of physicians and the challenges it poses.

This study surveyed 636 physicians across England to better understand limited EHR interoperability. Most participants reported that interoperability between NHS facilities was inadequate. Consequences of this included increased duration of hospital stays, lengthened consultation times, and more redundant diagnostic tests performed. Limited interoperability hindered communication between NHS workers and threatened care quality and patient safety. As more healthcare technologies are incorporated into the NHS, gaining greater insight from physicians is critical to finding solutions to address these problems.

## Introduction

Electronic health records (EHR) are commonplace in many healthcare settings and are often indispensable in care delivery. EHR replace paper-based medical charts to document patient clinical history, disease progression and medications, facilitate billing, and enable communication with other healthcare providers (1–3). However, their implementation in the preceding decades has been fraught with difficulty (3–6). In the English NHS, various national policies and local initiatives have hastened the technology’s introduction into clinical settings (4,7). However, this resulted in a patchwork of EHR systems being adopted, but with limited clinical data sharing capabilities between them (4,7–9). This contributed to considerable data fragmentation across various providers, suboptimal use of health data to improve overall care quality, and a largely inefficient and frustrating care seeking experience for patients (8,10).

Interoperability can facilitate effective care coordination, clinical decision support, and healthcare user satisfaction (11–13). However, the inability to easily access, modify, and share clinical information has been highlighted by many (14–18). For physicians and patients, poor EHR interoperability reportedly negatively impacted work productivity, created additional communication barriers between clinical teams, increased clinician burnout, necessitated time-consuming workarounds, and compromised patient safety (12–16,19–21). Poor interoperability was also noted to be detrimental to overall EHR data quality, as it contributed to patient data fragmentation (7,15,22).

While research examining EHR implementation is extensive, studies focussing on the interoperability of EHR systems are comparatively scarce (17,23–25). A Canadian study (2018) investigated what elements of interoperable EHR systems in emergency departments in Alberta were most useful to physicians. The authors found that most of the time spent on EHRs pertained to reviewing patient information useful for clinical decision making (26). Clinical settings utilising read-only interoperable EHR systems which lacked built-in patient management or clinical decision support tools, contributed to EHR disuse even when compared to those still reliant on paper-based processes. Altogether, the authors concluded that the clinical impact of using interoperable EHRs remained ‘poorly understood’ (26).

Another study explored the experiences of EHR use in primary care and community-based behavioural health settings in the US (27). The study highlighted that providers often resorted to various workarounds to compensate for a lack of EHR interoperability (27). These included the duplication of data entry and documentation, physically printing documents to scan or fax to share, relying on patients’ and clinicians’ recollection for information and using ‘freestanding’ tracking systems (*e.g.,* an Excel spreadsheet) (27). Another American study explored the facilitators and barriers to interoperability through interviews with hospital leaders, primary care providers, behavioural health providers, and regional health information exchange networks (23). The authors found that the expansion of HIT applications to suit differing needs but are otherwise not interoperable can contribute to data fragmentation and information overload for end-users (23). The resulting siloing of clinical information was recognised to potentially jeopardise the safety, quality, and efficiency of care (23).

To the best of our knowledge, few have quantified and mapped the prevalence of poor EHR interoperability and its perceived effects on quality and safety from physician perspectives (18,28). This study aimed to understand how physicians perceive a lack of interoperability impacts their day-to-day clinical activities, affects patient safety and changes the productivity of their work.

### Aims

The overall aims of this study are threefold:

1. To capture the perceptions of physicians regarding the current state of EHR interoperability;
2. To investigate how a lack of interoperability affects patient care and patient safety;
3. To estimate the effect of a lack of EHR interoperability on care efficiency and costs.

## Methods

### Study Population

Participants were practicing NHS doctors at different stages of training, ranging from trainees to consultants across both community and hospital-based settings. This included a wide range of specialties such as internal medicine, surgery, emergency medicine, anaesthesia, general practice, paediatrics, and psychiatry.

### Sampling

The calculated minimum sample size required for this study was 287 respondents. This was determined using Cochran’s Formula based on a desired 95% confidence level, 5% margin of error, an anticipated response rate of 25%, for a population size of 67,066 physicians currently working in the four main specialties most commonly associated with providing care for patients with chronic conditions (*i.e.,* internal medicine, surgery, emergency medicine, general practice (GP)) currently employed in the NHS, reported as of September 2020 (29). A more conservative anticipated response rate was selected due to the ongoing COVID-19 pandemic at the time this study was being conducted. However, this value is still consistent with similar estimates found in the literature regarding the use of web-based surveys (30–33). Convenience sampling and snowballing techniques were used.

### Participant Recruitment

The research team emailed existing contacts at Health Education England (HEE) deaneries and other relevant institutions (*e.g.,* Royal College of General Practitioners) with a request to circulate the study advertisements widely via e-mail with members in their immediate clinical networks who met the inclusion criteria, as well as to personal contacts in other clinical settings. Study advertisements were circulated amongst trainee physicians by local heads of departments or trainee representatives who have agreed with advertising the study in their healthcare facilities and via electronic newsletters. Reminders via e-mail and newsletters were sent out approximately every fortnight during the data collection period. No financial compensation was provided to participants. No follow-up assessments were held after data collection ended. Data collection lasted from June to October 2021.

### Description of Questionnaire

The questionnaire comprised 42 questions and was available in English only (**Appendix 1**). The content was developed based the existing literature and feedback from frontline NHS physicians. This was organised into four sections:

- Part 1: Basic participant demographic information
- Part 2: Physician EHR usage experience
- Part 3: Implications of EHR interoperability on patient safety and clinical care
- Part 4: Costs of healthcare resources accrued due to poor EHR interoperability in NHS hospitals.

The survey was piloted with four doctors (one GP, two surgical registrars, and one internal medicine trainee) and iteratively refined. The survey was hosted on Qualtrics, a web-based survey platform (34,35).

### Data Analysis

Descriptive statistics were calculated for survey respondents, with cross-tabulations of respondent characteristics against survey responses also performed. Response rates per question were calculated and the characteristics of respondents to each question were examined to identify any bias due to attrition over the survey course.

For questions exploring redundant diagnostic investigations performed, responses to individual types of tests (*e.g.,* FBC, urine dipstick, X-ray) were aggregated into ‘investigation-type’ categories (*e.g.,* blood-based, urine-based, radiological). An aggregated list of investigations is provided in **Appendix 2)**. When grouping responses within each ‘investigation-type’, the most frequent response for any constituent test was used to represent the maximum number of tests conducted per category.

Two-tailed ^2^ tests were performed to identify the relationship between participant characteristics of those who started the survey and completed it, with those participants who started the survey but did not complete the survey. All statistical analyses were performed using Stata 13.1.

## Results

### Participant Characteristics

A total of 636 NHS doctors participated in the survey, of which 218 (34.3%) have completed it in its entirety. A full description of the respondents is provided in **Table 1**. Of those who responded, 47.4% were females, 37.2% were aged between 30-39. London (n=155, 28%) and Northwest England (n=131, 23.7%) received the greatest number of responses, and 48.1% were working in academic hospital settings. GPs comprised the largest clinical training group amongst participants (n=266, 44.1%). Participants who completed the survey were typically older in age, held more senior clinical roles, worked in internal medicine and had no formal EHR training **(Appendix 3)**.

**Table 1:** Characteristics of study participants. Percentages expressed in the ‘Missing’ rows are based on the total number of study participants.

| Characteristics | n | Out of total number of participants, (%) | Of those who responded, (%) |
| --- | --- | --- | --- |
| **Gender** |  |  |  |
| Female | 282 | 44.3% | 47.4% |
| Male | 313 | 49.2% | 52.6% |
| Missing | 41 | 6.5% |  |
| **Age band (years)** |  |  |  |
| Under 30 | 122 | 19.2% | 20.2% |
| 30-39 | 224 | 35.2% | 37.2% |
| 40-49 | 145 | 22.8% | 24.1% |
| 50-59 | 84 | 13.2% | 13.9% |
| 60+ | 28 | 4.4% | 4.6% |
| Missing | 33 | 5.2% |  |
| **Clinical role** |  |  |  |
| Foundation year (FY1-2) | 59 | 9.3% | 9.8% |
| Senior house officers (ST1-2) | 98 | 15.4% | 16.3% |
| Registrar/Senior registrar (ST3+) | 180 | 28.3% | 29.9% |
| Consultant/GP | 266 | 41.8% | 44.1% |
| Missing | 33 | 5.2% |  |
| **Location of practice** |  |  |  |
| East of England | 52 | 8.2% | 9.4% |
| London | 155 | 24.4% | 28% |
| Midlands | 54 | 8.5% | 9.8% |
| North East and Yorkshire | 91 | 14.3% | 16.4% |
| North West | 131 | 20.6% | 23.7% |
| South East | 30 | 4.7% | 5.4% |
| South West | 41 | 6.5% | 7.4% |
| Missing | 82 | 12.9% |  |
| **Medical specialty training** |  |  |  |
| Internal medicine and subspecialties | 226 | 35.5% | 37.7% |
| Surgery and subspecialties | 123 | 19.3% | 20.5% |
| A&E | 56 | 8.8% | 9.3% |
| Anaesthesia | 48 | 7.6% | 8% |
| GP/Family medicine | 50 | 7.9% | 8.3% |
| Paediatrics | 43 | 6.8% | 7.2% |
| Psychiatry | 41 | 6.5% | 6.8% |
| Other | 13 | 2% | 2.2% |
| Missing | 36 | 5.7% |  |
| **Type of healthcare facility** |  |  |  |
| Academic/teaching hospital | 290 | 45.6% | 48.1% |
| District general/community hospital | 244 | 38.4% | 40.5% |
| GP practice | 42 | 6.6% | 7% |
| Other | 27 | 4.3% | 4.5% |
| Missing | 33 | 5.2% |  |
| **Number of organisations interacting with** |  |  |  |
| None | 7 | 1.1% | 1.5% |
| 1-2 | 58 | 9.1% | 12% |
| 3-4 | 144 | 22.6% | 29.8% |
| 5+ | 275 | 43.2% | 56.8% |
| Missing | 152 | 23.9% |  |
| **Received formal EHR training** |  |  |  |
| Yes | 256 | 40.3% | 56.5% |
| No | 197 | 30% | 43.5% |
| Missing | 183 | 28.8% |  |
| **Self-reported EHR proficiency level** |  |  |  |
| Beginner | 23 | 3.62% | 5% |
| Moderate | 235 | 37% | 50.7% |
| Advanced | 167 | 26.3% | 36% |
| Expert | 39 | 6.1% | 8.4% |
| Missing | 172 | 27% |  |
| **Years of experience using EHR** |  |  |  |
| Less than 1 year | 25 | 3.9% | 5.4% |
| 1-2 years | 101 | 16% | 22% |
| 3-5 years | 138 | 21.7% | 30% |
| 6-10 years | 115 | 18.1% | 25% |
| More than 10 years | 83 | 13.1% | 18% |
| **Number of clinical sessions per week** |  |  |  |
| None | 1 | 0.2% | 2.3% |
| 1-2 | 7 | 1.1% | 16.3% |
| 3-4 | 10 | 1.6% | 23.3% |
| 5+ | 25 | 3.9% | 58.1% |
| Missing | 593 | 93.2% |  |
| **Frequency of EHR use** |  |  |  |
| Less than once a month | 2 | 0.3% | 0.4% |
| At least once a month | 7 | 1.1% | 1.5% |
| At least once a week | 31 | 4.9% | 6.7% |
| Every day | 424 | 66.7% | 91.4% |
| Missing | 172 | 27% |  |

### Perceptions of Healthcare Providers Regarding the Current State of EHR Interoperability

#### Interoperability-related EHR functions currently available and in use

Recognising that what interoperability-related EHR functions (*i.e.,* functions which involve the input and transfer of health data between two or more EHR elements or users) are available to NHS doctors may not necessarily align with what are routinely used, participants were asked to identify which functions were present and in common use at their workplace **(Table 2)**. The three most commonly available functions highlighted by respondents were (1) retrieval of patient’s previous health information (n=429/461, 93.1%), (2) inputting orders for investigations and medications (n=411/461, 89.2%), and (3) planning patient disposition and discharges (n=329/461, 71.4%). The most frequently used functions reported were (1) retrieval of patient’s previous health information (n=291/461, 63.1%), (2) inputting orders for investigations and medications (n=261/461, 56.6%), and communicating with other healthcare professionals (n=163/461, 35.4%).

**Table 2:**
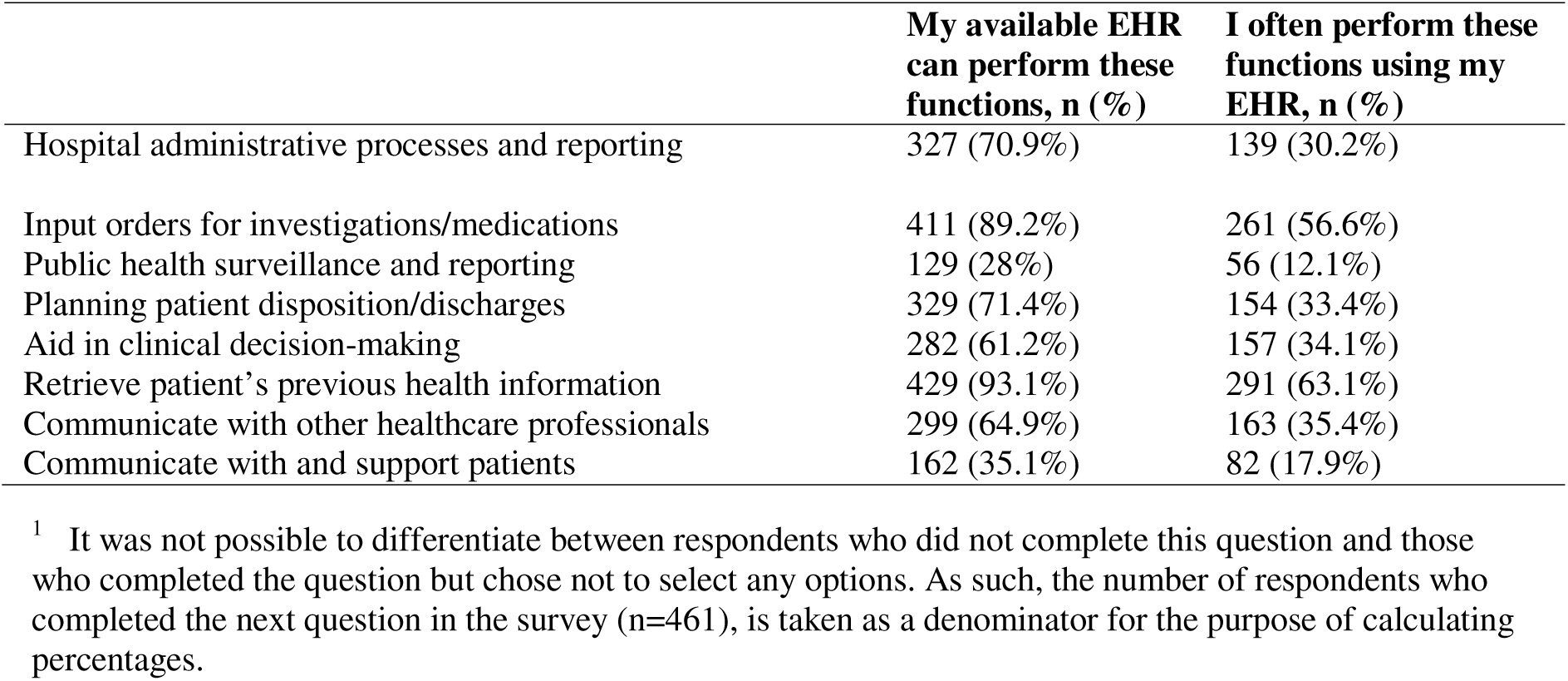
EHR functions commonly available and in use by NHS doctors.

|  | My available EHR<br>can perform these<br>functions, n (%) | I often perform these<br>functions using my<br>EHR, n (%) |
| --- | --- | --- |
| Hospital administrative processes and reporting | 327 (70.9%) | 139 (30.2%) |
| Input orders for investigations/medications | 411 (89.2%) | 261 (56.6%) |
| Public health surveillance and reporting | 129 (28%) | 56 (12.1%) |
| Planning patient disposition/discharges | 329 (71.4%) | 154 (33.4%) |
| Aid in clinical decision-making | 282 (61.2%) | 157 (34.1%) |
| Retrieve patient's previous health information | 429 (93.1%) | 291 (63.1%) |
| Communicate with other healthcare professionals | 299 (64.9%) | 163 (35.4%) |
| Communicate with and support patients | 162 (35.1%) | 82 (17.9%) |

#### Directionality of interoperability present in existing EHR systems

Most respondents reported that they can view clinical information inputted by other healthcare providers within their own healthcare setting or facility (n=418/461, 90.7%). However, visibility of clinical information outside of their immediate healthcare setting (n=175/460, 38%) and the ability for external healthcare providers to see their inputted data (n=74/457, 16.2%), were markedly lower.

Most respondents stated that they cannot edit clinical information within participants’ healthcare setting (n=225/460, 48.9%) and that from external healthcare providers (n=381/452, 84.3%). Conversely, clinical information in the participant’s hospital or clinic is typically not viewable and editable by most external healthcare providers (n=309/456, 67.8%). Of note was the increase in the number of ‘I do not know’ responses corresponding with EHR interactions of increasing complexity.

### Impact of EHR interoperability on patient care

Most participants (n=396/413, 95.9%) reported that they have experienced difficulties retrieving clinical information from EHR systems. Of these, a quarter (n=100, 24.2%) stated that this occurred most of the time or always. Most respondents reported that poor EHR interoperability negatively impacted their day-to-day clinical workflow (n=386/412, 93.7%), of which 127 (n=127, 30.8%) stated that this occurred most of their time or always. Similarly, 81.5% (n=335/411) of respondents reported that they felt poor EHR interoperability posed a potential risk patient safety, with 19.2% (n=79) describing it as a risk most of the time or always. Most clinicians reported that poor EHR interoperability negatively affected their ability to share clinical information with other healthcare professionals (n=450/411, 91.8%). Similarly, 352 of 409 respondents (86.1%) described it as being detrimental to them communicating with patients and caregivers.

### Impact on EHR Data Visibility and Tasks

Three problems associated with data visibility and completion of EHR tasks most frequently identified by respondents were: (1) Difficulty retrieving patient information available in another healthcare (n=300, 83.6%), (2) Difficulty accessing patient information even when you know that information is available locally within the system (n= 217, 60.5%), and (3) Difficulty conveying clinical information for another healthcare professional (n=216, 60.2%).

### How Interoperability Affects Patient Care Safety

When asked to rate their overall experience of EHRs and interoperability in the current workplace, participants reported largely positive (‘Good’, n=124, 30%) or neutral experiences (‘Neutral’, n=117, 28.3%). ‘Bad’ (n=90, 21.7%), ‘Very bad’ (n=55, 13.3%), and ‘Very good’ (n=28, 6.8%) comprised the remainder of responses received. A large proportion of negative experiences (Very bad and Bad) was reported by those having a lower self-reported EHR proficiency. Conversely, GPs and doctors practicing in non-secondary care centres, tended to report comparatively positive experiences with EHR interoperability in contrast to other specialties. The reported experiences cross tabulated against the various participant characteristics are shown in **Figure 1**.

**Figure 1:**
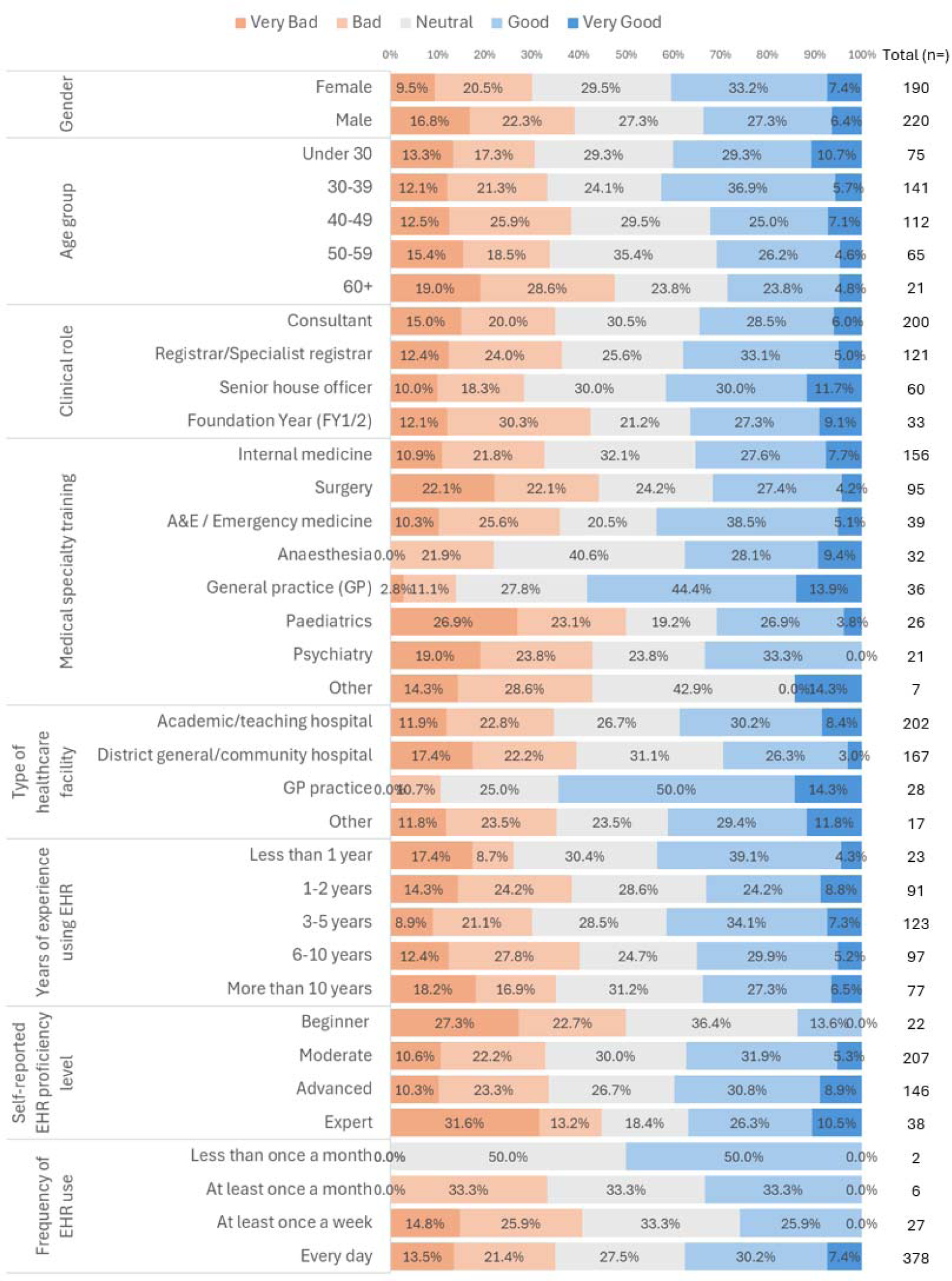
Breakdown of characteristics of doctors reporting experiences of EHR interoperability at their workplace.

Out of 413 responses received, most participants (n=366, 88.6%) indicated they believed that the lack of EHR interoperability does or may pose a risk to patient safety, with 224 (52.2%) providing a ‘Yes’ response. The perceived risk to patient safety is most prevalent amongst doctors in specialties based in secondary care centres (*e.g.,* surgery, A&E, and paediatrics), and those have had less than a year’s experience using EHRs. For a breakdown of the responses by participant characteristics, please see **Figure 2**.

**Figure 2:**
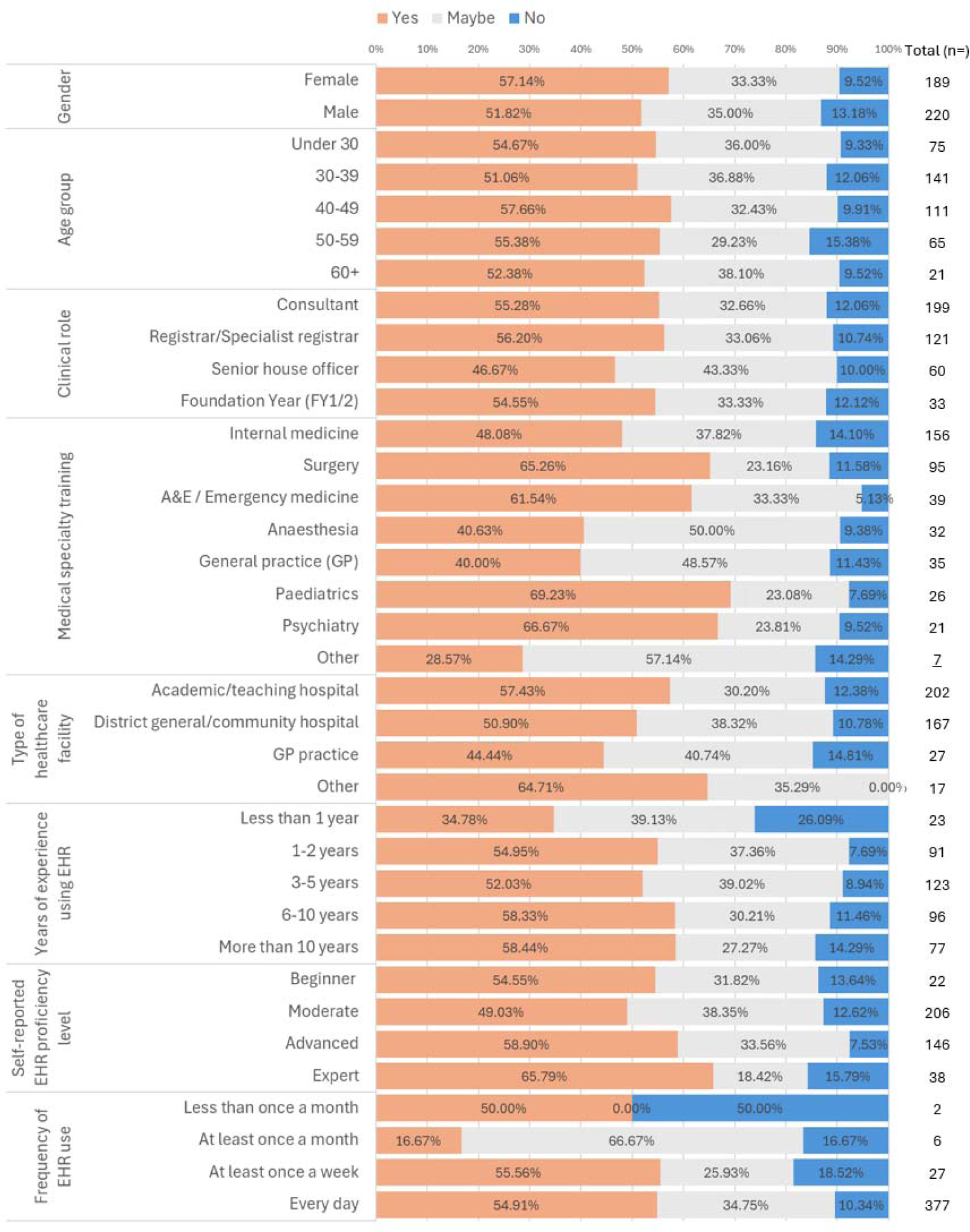
Doctor perceptions of whether limited EHR interoperability poses a patient safety risk.

### Mapping impact of the lack of EHR interoperability along pathway of care

To identify where along the clinical care pathway the lack of EHR interoperability typically occurs, participants were asked when they find it most impeding to their clinical work during a routine clinical shift. Out of the total number of study participants, only 310 (48.7%) completed this question **(Table 6)**. The most common response was when receiving patients from a secondary or tertiary healthcare facility (n=224, 72.3%), followed by medication reconciliation (n=178, 57.4%), and transferring patients to another healthcare facility (n=175, 56.5%).

**Table 3:**
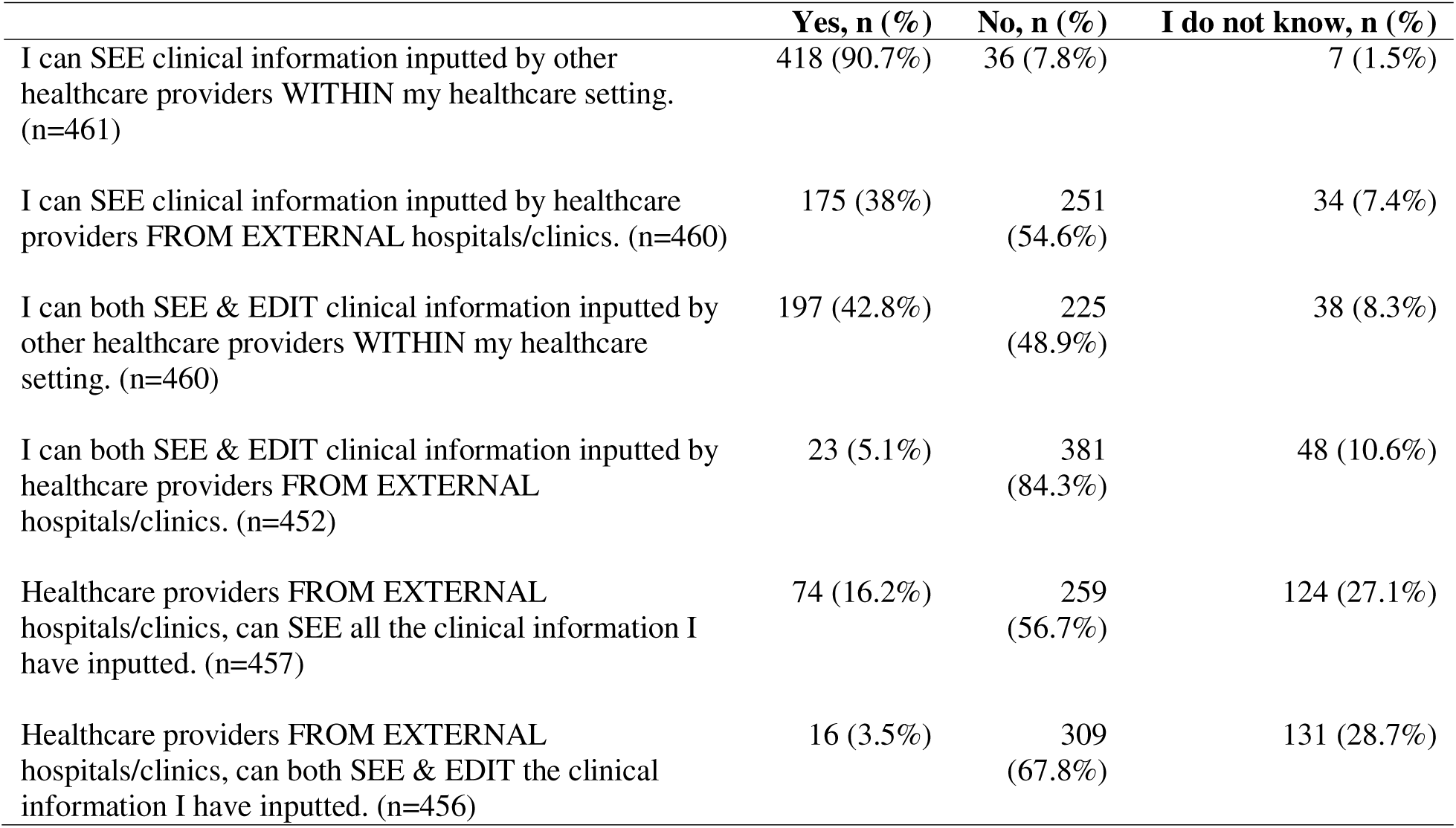
Directionality of interoperability of existing EHR systems.

**Table 4:** Impact of interoperability on patient care, safety, and clinical workflow.

|  | Always, n (%) | Most of the time, n (%) | About half the time, n (%) | Sometimes, n (%) | Never, n (%) |
| --- | --- | --- | --- | --- | --- |
| Difficulty accessing and retrieving clinical information through the EHR systems currently in use. (n= 413) | 38 (9.2%) | 62 (15%) | 64 (15.5%) | 232 (56.2%) | 17 (4.1%) |
| Difficulty with accessing and retrieving this clinical information negatively affects my day-to-day clinical workflow. (n=412) | 55 (13.4%) | 72 (17.5%) | 56 (13.6%) | 203 (49.3%) | 26 (6.3%) |
| Difficulty with accessing and retrieving this clinical information poses a potential risk to patient safety during my routine shifts in the hospital/clinic. (n=411) | 30 (7.3%) | 49 (11.9%) | 27 (6.6%) | 229 (55.7%) | 76 (18.5%) |
| Difficulty with accessing and retrieving this clinical information negatively impacts my ability to share clinical information with other healthcare professionals. (n=411) | 46 (11.2%) | 72 (17.5%) | 55 (13.4%) | 198 (48.2%) | 40 (9.7%) |
| Difficulty with accessing and retrieving this clinical information negatively impacts my ability to share clinical information with my patients and/or their caregivers. (n=409) | 47 (11.5%) | 56 (13.7%) | 46 (11.3%) | 203 (49.6%) | 57 (13.9%) |
| Thinking about your patients' expectations regarding the accessibility of their health records, do you feel that the EHR systems you currently use allow you to meet these expectations? (n=408) | 21 (5.2%) | 76 (18.6%) | 70 (17.2%) | 165 (40.4%) | 76 (18.6%) |

**Table 5:**
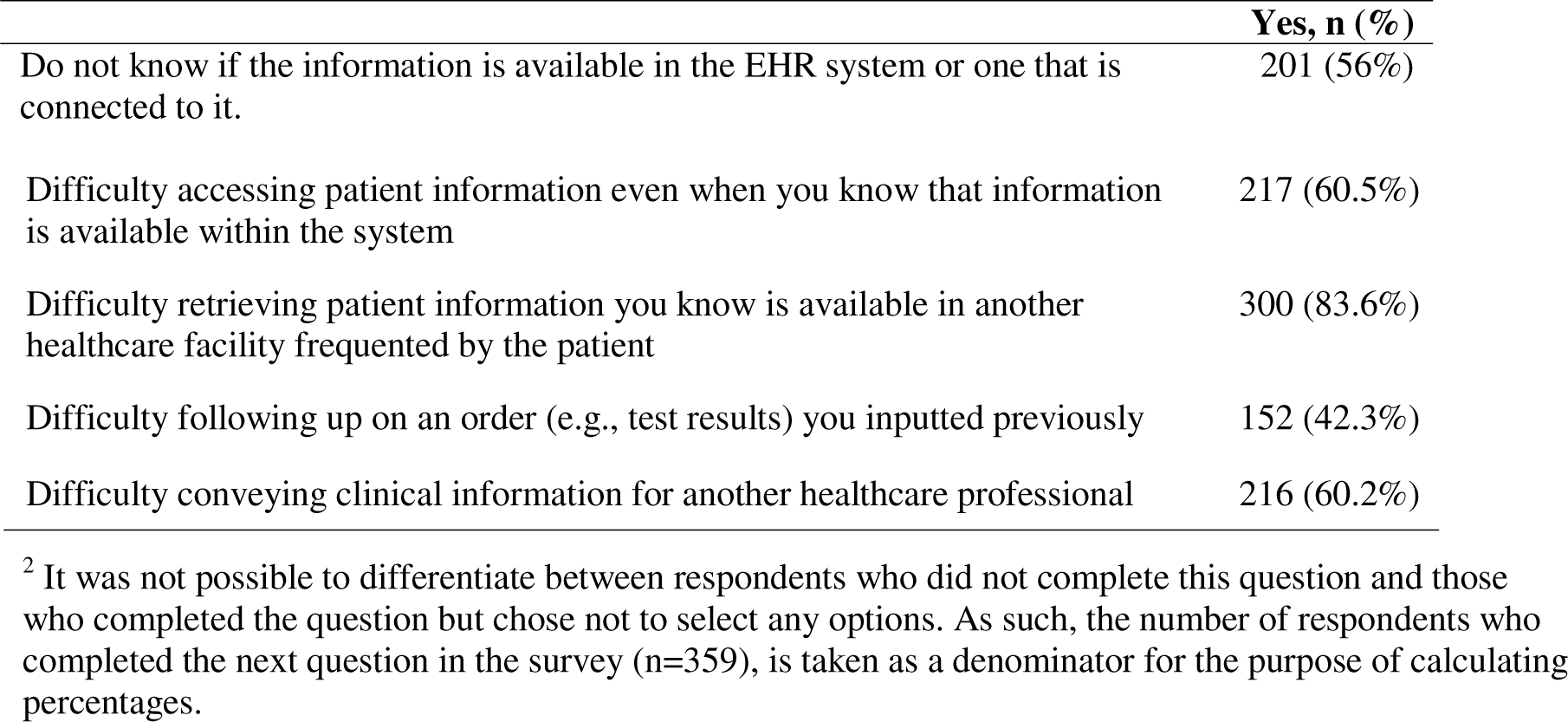
Difficulties with EHR data availability and tasks due to poor interoperability.

|  | Yes, n (%) |
| --- | --- |
| Do not know if the information is available in the EHR system or one that is connected to it. | 201 (56%) |

<sup>2</sup> It was not possible to differentiate between respondents who did not complete this question and those who completed the question but chose not to select any options. As such, the number of respondents who completed the next question in the survey (n=359), is taken as a denominator for the purpose of calculating percentages.

|  |  |
| --- | --- |
| Difficulty accessing patient information even when you know that information is available within the system | 217 (60.5%) |
| Difficulty retrieving patient information you know is available in another healthcare facility frequented by the patient | 300 (83.6%) |
| Difficulty following up on an order (e.g., test results) you inputted previously | 152 (42.3%) |
| Difficulty conveying clinical information for another healthcare professional | 216 (60.2%) |

**Table 6:** When is the lack of interoperable EHRs most impeding to your clinical work during a routine clinical shift?

|  | Yes, n (%) | No, n (%) |
| --- | --- | --- |
| Admitting a new patient from the community | 152 (49%) | 158 (51%) |
| Receiving a patient from a secondary/tertiary healthcare facility | 224 (72.3%) | 86 (27.7%) |
| During handover of patients from other members within my immediate clinical team | 51 (16.5%) | 259 (83.5%) |
| During handover of a patient from another clinical team/clinician within my hospital | 71 (22.9%) | 239 (77.1%) |
| Following up on an order from another clinical team/clinician in my hospital | 102 (32.9%) | 208 (67.1%) |
| Discharging patient from my hospital back into the community | 102 (32.9%) | 208 (67.1%) |
| Transferring a patient from my hospital to another secondary/tertiary healthcare facility | 175 (56.5%) | 135 (43.6%) |
| Medication reconciliation | 178 (57.4%) | 132 (42.6%) |
| Other | 28 (9%) | 282 (91%) |

### Care Inefficiencies and Associated Healthcare Costs Incurred

To gain a better understanding of the impact on physician productivity resulting from poor EHR interoperability, participants were asked whether they felt poor EHR interoperability has negatively impacted their workflow in terms of: (1) repeat diagnostic investigations, (2) prolonged length of stay in hospital, and (3) prolonged clinic preparation and consultation times. Participants who responded ‘Yes’ were then asked to quantify their response for each category.

#### Repeat diagnostic investigations

For repeat diagnostic investigations, participant responses were aggregated into investigation type categories (**Table 7)**. Responses for each investigation type are detailed in **Appendix 2**.

**Table 7:** Repeat diagnostic investigations performed because of poor EHR interoperability as reported by 289 respondents who completed the question.

|  | Bloodwork, n (%) | Blood Gases, n (%) | Urine-based, n (%) | Infectious Panel, n (%) | Radiology, n (%) |
| --- | --- | --- | --- | --- | --- |
| Never | 108 (37.4%) | 207 (76.1%) | 125 (46.8%) | 21 (8%) | 123 (42.9%) |
| Once a week | 114 (39.5%) | 48 (17.7%) | 87 (32.6%) | 33 (12.5%) | 125 (43.6%) |
| 2-3 times a week | 35 (12.1%) | 11 (4%) | 30 (11.2%) | 13 (4.9%) | 25 (8.7%) |
| 4-6 times a week | 15 (5.2%) | 3 (1.1%) | 12 (4.5%) | 3 (1.1%) | 5 (1.7%) |
| Daily | 17 (5.9%) | 3 (1.1%) | 13 (4.9%) | 194 (73.5%) | 9 (3.1%) |

Infectious panels were reported to be the most repeated on a daily basis due to poor interoperability (n=194, 73.5%). Radiological investigations were the second most repeated, with physicians reportedly needing to do so once per week (n=125, 43.6%). This is followed by bloodwork (n=114, 39.5%) and urine-based investigations (n=87, 32.6%), both of which are require repeating typically on a weekly basis.

#### Prolonged length of stay (PLOS) in hospital

When questioned as to whether problems sharing or retrieving clinical information via EHRs with limited interoperability resulted in keeping patients in hospital longer, 143 out of 222 (64.4%) respondents responded ‘Yes’. When asked to quantify the extent of delay for patients where limited EHR interoperability contributed to a problem, 105 (42.9%) participants reported that issues with EHR interoperability often resulted in several hours in delays in discharging patients, 53 (21.6%) reported that it typically caused 1-night additional stay in hospital, 52 (21.2%) saw no delays caused, and 35 (13.2%) caused 2+ nights of additional hospital stay.

Delays of several hours were reported most frequently from surgical subspecialties (n=42, 51.9%), medicine subspecialties (n=21, 29.2%), and A&E (n=17, 51.5%) **(Table 8)**. Delays causing one additional night stay were most reported by medicine subspecialties (n=25, 34.7%), followed by A&E (n=9, 27.3%), and surgical subspecialties (n=9, 11.1%).

**Table 8:** Cross tabulation of delays in hospital vs. specialty training. Percentages expressed are for values per row.

|  | No delays,<br>n (%) | Several hours<br>delay, n (%) | 1-night additional<br>stay in hospital, n (%) | 2+ nights additional<br>stay in hospital, n (%) |
| --- | --- | --- | --- | --- |
| Internal medicine | 6 (8.3%) | 21 (29.2%) | 25 (34.7%) | 20 (27.8%) |
| Surgery | 21 (25.9%) | 42 (51.9%) | 9 (11.1%) | 9 (11.1%) |
| A&E | 5 (15.2%) | 17 (51.5%) | 9 (27.3%) | 2 (6.1%) |
| Anaesthesia | 3 (18.8%) | 9 (56.3%) | 3 (18.8%) | 1 (6.3%) |
| Paediatrics | 8 (36.4%) | 11 (50%) | 2 (9.1%) | 1 (4.6%) |
| Psychiatry | 8 (53.3%) | 1 (6.7%) | 4 (26.7%) | 2 (13.3%) |
| Other | 1 (20%) | 3 (60%) | 1 (20%) | 0 (0%) |

#### Prolonged clinic consultation times

Most participants stated that interoperability problems have necessitated extra time during routine clinic consultations **(Table 9)**, both when preparing for (n=209, 95.9%) as well as during consultations themselves (n=211, 96.8%). Most participants required between an extra 15-30 minutes of preparation time (n=72, 33%) and another 15-30 minutes of consultation time during a routine day of clinic consultations (n=71, 32.6%).

**Table 9:** Prolonged clinic times due to issues of EHR interoperability amongst participants who responded (preparation and during the consultation).

|  | Preparing for consultations, n (%) | During consultations, n (%) |
| --- | --- | --- |
| No extra time needed | 9 (4%) | 7 (3.2%) |
| Less than 5 minutes | 18 (8.3%) | 22 (10.1%) |
| 5-15 minutes | 56 (25.7%) | 64 (29.4%) |
| 15-30 minutes | 72 (33%) | 71 (32.6%) |
| 30-60 minutes | 45 (20.6%) | 43 (19.7%) |
| More than an hour | 18 (8.3%) | 11 (5.1%) |

## Discussion

### Summary of Principal Results

This survey of 636 NHS doctors in England reveals provides an important insight into how electronic health record interoperability, or a lack thereof, affects their day-to-day practice.

EHRs are widely used by clinicians to retrieve clinical information about their patients, and also to input information into clinical systems, including for the ordering of investigations and medications. Further, a significant minority of doctors report using EHRs to communicate with other healthcare colleagues.

While most doctors are able to view patient information in their own hospital EHRs (91%), far fewer are able to edit this information (43%), likely reflecting constraints on editing medical records written by others. Many respondents reported being able to view information from other healthcare organisations (38%) but did not believe other organisations were able to view the information they had inputted (16%). This finding potentially indicates either a lack of reciprocity in EHR interoperability, or a lack of awareness of the interoperability capabilities of providers with whom clinicians regularly share patients. This latter point is supported by increasing numbers of ‘don’t know’ responses to questions pertaining to what was possible at other providers than the respondent’s own. When attempting to map out where issues with limited EHR interoperability typically occur along a patient’s clinical pathway, participants largely highlighted this manifesting during the transition of care between two separate NHS settings, as well as when doctors are completing medication reconciliation.

Almost all participants reported experiencing some form of difficulty in accessing clinical information due to poor EHR interoperability. Unsurprisingly, many stated that this was detrimental to their clinical workflow and impaired their communication with patients and other healthcare providers. In terms of current experiences of interoperability at their workplace, negative views were more prevalent amongst doctors who practiced in specialties based in secondary care settings as well as those with less than one year of EHR experience and lower self-reported EHR proficiency levels. Regarding perceived impact on patient safety overall, most doctors reported that limited EHR interoperability they routinely experienced does pose some form of risk. This finding consistently outnumbered those who did not feel it was so across all participant characteristics.

However, the impact of lack of EHR interoperability on care efficiency was mixed. Infection panels were repeated by many respondents on a daily basis, with other redundant investigations being done so typically weekly. Most reported that poor EHR interoperability led to extended hospital length of stay and prolonged consultation times, though responses quantifying delays varied between specialties (*e.g.,* surgeons tended to report shorter delays than internists).

### Comparison with Prior Work

To better understand EHR systems used across the UK, Warren *et al.,* utilised national-level administrative data to explore the spatial distribution of EHR systems in relation to their use when patients seek care between NHS Trusts (7). The authors found that although a total of 21 different EHR systems were in use and three vendor’s systems made up the majority, most hospitals did not have robust standardised means of electronic clinical data sharing. Even when patients sought care between two trusts which utilised EHRs from the same vendor, only 0.6% of patients were able to take advantage of the greater interoperability possible (7). The authors highlighted that while 20 pairs of Trusts were found to routinely share the same cohort of patients, only one pair utilised EHR systems which were from the same vendor and allowed for some form of interoperability (7). Though this study was valuable in identifying macro-level trends, the secondary data used did not allow for more granular insight into how poor interoperability materialises along a patient’s care pathway.

Investigating the amount of additional healthcare expenditure because of poor interoperability has been explored. For example, Stewart *et al.,* examined whether poor interoperability was associated with increases in duplicate medical tests conducted in patients with congenital heart disease being shared between nearby two hospitals with no EHR interoperability in Boston, Massachusetts (36). Of the 27 patient records that showed duplicate investigations performed, 17 were not clinically indicated and half had more than one test duplicated (36). The majority of these were conducted during admission from outpatient clinics, with authors speculating that this was likely due to the delays or difficulties encountered during the transfer of test results (36). An estimated US$ 1255 was thus accrued to all the study patients because of duplicated investigations (US$ 14.56 per patient). While this sum was modest, the savings from minimising duplicative efforts is likely substantial (36). However, this study’s generalisability is limited as it pertained only to a highly specialised patient group and used a narrow definition of ‘duplicate test’ (36).

In a similar attempt to explore the potential cost benefits resultant from increased interoperability, Meyer *et al.,* measured the difference in perceived and actual time spent by nurses completing clinical task orders when using EHR systems with and without interoperability in Geneva-based hospitals (37). An EHR system used in one hospital without interoperability was compared to one introduced at another hospital which had interoperability features available. The authors found that interoperability between the various systems saved on average 26 seconds per order entry, allowing the systems to be utilised approximately three times as fast (37). When extrapolated with the wages for nurses at approximately US$ 0.73 per minute and roughly 20,000 laboratory orders generated monthly, the authors estimated that incorporating interoperability between the various HIT systems contributed on average, US$ 6325 of work-time equivalent savings per month (37).

### Strengths and Limitations

This is the first study exploring challenges posed to NHS physicians when using EHRs with varying levels of interoperability, ascertaining where interoperability-related issues arise on clinical pathways, and examining the impact to productivity in the NHS. This study comprised a relatively large sample size, received responses from physicians across England, and includes participants from hospital and community-based settings. The survey itself is methodologically robust, with the content derived from a systematic review of the wider literature and subsequently piloted with four physicians of varying clinical experience from three different specialties.

Limitations are largely inherent to survey studies themselves. Given that the findings are self-reported, there is a risk of self-selection and recall bias. While stratified purposive sampling was initially planned to ensure proportionate representation of physicians from different backgrounds, convenience sampling was eventually used due to practical considerations during the ongoing pandemic circumstances. Another limitation was the relatively high attrition rate amongst certain subgroups to finishing the study.

For example, the small number of responses from Foundation Year doctors at the beginning of the survey (n=59), dropping to a handful by the end (n=8), made an analysis of their responses a notable challenge. Given this subgroup tends to interact with EHR more senior staff, the lack of responses from them contribute to a loss of granularity in the overall data captured.

### Implications for Practice, Policy, and Further Research

NHS physicians largely reaffirmed that poor EHR interoperability is a widespread problem across the health system which negatively impacts their ability to deliver safe and efficient care.

GPs were the only standout group amongst the study cohort who reported comparatively positive experiences with EHR interoperability. This is potentially due to GPs generally having greater control over the EHR systems in use at their clinics and the less frequent need to interact with other clinical teams to facilitate routine patient care. However, given the high number of responses highlighting interoperability-related issues when receiving patients from secondary care (n=224), this suggests that problems with inter-organisational EHR interoperability are more pronounced than that within primary care settings themselves. Together, this may have contributed to mitigating some of the overall negative experiences perceived by GPs compared to doctors in secondary care.

The variation in negative views from doctors across the levels of self-reported EHR proficiency also pints to a potential gap in EHR skills for young trainees entering their first clinical workplace. Once junior doctors are familiarised with the EHRs available, the slight reduction of negative views suggest that greater experience may have helped them overcome some of the initial perceived systems shortcomings (*e.g.,* workflow inconveniences). Those who eventually become ‘experts’ with their EHRs are likely to be more vocal about the negative experiences surrounding interoperability issues routinely encountered. This trend is similar regarding the perceived risk to patient safety – those who have had less time using EHRs have yet to recognise it as a threat to patient safety compared to more seasoned users. Nonetheless, this did not alter the prevailing perception amongst most doctors that current levels of EHR interoperability remain inherently unsafe.

Regarding impact on clinical tasks, responses largely indicated the inability to view results of recently conducted investigations and difficulty in communicating with other health professionals. The mismatch in responses quantifying the severity of interoperability-related challenges versus the perception of risks to care, might be explained by limited interoperability causing poor end-user experiences, making it easier for errors to occur and indirectly causing harm.

Deriving an estimate of the impact of poor interoperability on productivity proved especially challenging. Nonetheless, respondents were able to highlight some notable areas of concern. For example, physicians reported that radiological investigations were the second most repeated type of investigations due to poor interoperability, with this occurring typically once a week. With investigations such as MRIs having a unit cost averaging £161.54, these extra investigations have the potential to add up to more significant costs when extrapolated across the NHS (38,39). While most participants noted that interoperability-related discharge delays were common, surgical subspecialties typically kept patients for several more hours, while medical subspecialties reported longer delays, typically one additional night, and up to 2-3 nights. Costs of delays lasting ‘several hours’ are difficult to establish given the imprecise definition, the opportunity costs based on bed location (*e.g.,* A&E versus wards), and the professionals involved. For reference, a non-elective short stay (*i.e.,* two days or less) is estimated to be on average £801per unit cost (38).

Recommendations to mitigate some of these challenges are gradually becoming available. The NHS Interoperability Toolkit, first released in 2010 and recently updated in March 2023, comprises a collection of frameworks, guidance material, and technical reference documentation to streamline validation processes, lower costs, and provide set interoperability standards for vendors (40,41). Policy-based approaches, such as those suggested by Zhang *et al.,* proposed having regulatory bodies (*e.g.,* Care Quality Commission (CQC) and NHSX) promote, enforce, and push for systemic changes which have proven difficult to do with the currently fragmented HIT procurement strategies between trusts and the lack of business incentives for system vendors to do so themselves (42). While regulatory bodies do not have the authority to dictate HIT policies directly, greater regulatory involvement would give a stronger sense of collective strategic direction in realising a more integrated network of EHRs.

Future research should capture the perceptions of EHR interoperability from other healthcare workers (*e.g.,* nurses, pharmacists, allied health workers), ascertain how problems arise along clinical pathways, and measure the impact on patient care and safety using standardised key performance indicators. Given that other NHS healthcare professionals such as nurses, pharmacists, and allied health professionals, may be similarly affected by limited EHR interoperability, there is a clear imperative to explore these perspectives. Research examining EHR user experience, technical aspects of interoperability, or tangential areas such as implementation sciences and economic analyses can advance the understanding of the value posed by interoperable EHRs (43).

## Conclusion

In the English NHS, EHR interoperability is minimal, sporadic, and poses a multitude of practical workflow, communication, and potential safety challenges for frontline NHS doctors. While there is a broad consensus regarding its detrimental effects, the perceived impact of poor interoperability is not felt uniformly across the NHS. Certain physician characteristics such as specialty or years of experience using EHRs, influence their perception of interoperability and how they believed it could impact patient care. Poor interoperability was reported to necessitate repeat diagnostics, lengthen hospital stays, and prolong clinic consultations, though the related burdens were not perceived to be severe. However, when extrapolated across the whole of the NHS, these seemingly less significant hinderances would likely culminate in substantial inefficiencies and costs.

This study illuminates the perceptions of poor interoperability in practice from users themselves. As the first of its kind in the UK attempting to ‘take the pulse’ of how robust health information exchanges are implemented across various healthcare settings, this study sets the stage for future efforts to more thoroughly investigate the extent the problem that poor EHR interoperability poses. Future efforts at overhauling EHR interoperability must incorporate technical as well as policy-based solutions which closely reflect the practical needs, concerns, and feedback of end-users.

## Data Availability

All data produced in the present study are available upon reasonable request to the authors.

## Ethical Approval

Overall ethical approval for this project was granted by the Imperial College Research Ethics Committee (ICREC) (Reference no. 20IC5906). This is a dedicated ethics oversight body at Imperial College London for all health-related research involving human participants.

## Abbreviations

API: application programming interface
CMS: US Centers for Medicare & Medicaid Services
CPOE: computerised provider/physician order entry
EHR: electronic health records
FHIR: Fast Healthcare Interoperability Resources
GP: general practitioner
HIT: health information technology
HL7: Health Level 7
NHS: National Health Service
SHO: senior house officer
PLOS: patient length of stay

