## Appendices 1-7 for "Physician experiences of electronic health records interoperability and its practical impact on care delivery in the English NHS: A cross-sectional survey study"

### Appendix 1

Electronic Health Records, Interoperability, and Patient Safety - An NHS Clinician Survey

Survey Flow

Standard: PIS & Consent (2 Questions)

Block: Part 1: Basic Participant Information (8 Questions)

Standard: Part 2: Clinician EHR User Experience (7 Questions)

Standard: Part 3: Implications of EHR Interoperability on Patient Safety & Clinical Care (12 Questions)

Standard: Part 4: Costs of Healthcare Resources Used Due to Poor EHR Interoperability (13 Questions)

| Page Break |
| --- |

Start of Block: PIS & Consent

Q37 You are invited to participate in a research study exploring how electronic health records (EHR) and interoperability can impact patient safety and costs in the NHS. This study is being conducted by Dr. Ana Luisa Neves and her research team from the Patient Safety Translational Research Centre (PSTRC), Institute for Global Health Innovation (IGHI) at Imperial College London. 

To participate, you must be a qualified doctor currently practicing in NHS England, be over the age of 18, and be able to communicate in English. Participation in this study is entirely voluntary. If you agree to participate in this study, you will be completing an online questionnaire requiring approximately 10 minutes of your time. This can be done either via a traditional computer or smartphone. Whilst participating in this study may not benefit you directly, but it will help us better understand how doctors perceive the need for EHR interoperability and how it affects patient safety during routine clinical practice. This will be used to help inform future healthcare policy makers on how to better incorporate health informatics systems to meet the evolving needs of healthcare providers and patients alike. You may skip any questions you don’t want to answer and you may end the questionnaire at anytime. The information you will share with us, if you participate in this study, will be kept completely anonymous and confidential.

All responses collected from this study will be kept on an encrypted server belonging to the Imperial College Big Data Analytical Unit (BDAU) located on the 10th floor at St. Mary's Hospital Paddington. This will be stored for up to 10 years after the conclusion of this study and may be used for future research projects. Only the Principal Investigator and members of the research team will be able to access the collected data. No one else at the PSTRC, IGHI, or Imperial College London will be able to see your responses or even know whether you participated in this study. Study findings will be presented only in aggregate/summary form for publication in peer-reviewed journals. No identifiable information would be used in any report.

Q38 By selecting "I agree" and participating in this study, you are consenting to the conditions described above.

I agree (1)

I do not agree (2)

Skip To: End of Survey If Q38 = I do not agree

End of Block: PIS & Consent

Start of Block: Part 1: Basic Participant Information

Q1 Please select your gender:

Female (1)

Male (2)

Other (3)

Prefer not to answer (4)

Q2 Please select your age:

Under 30 (1)

30-39 (2)

40-49 (3)

50-59 (4)

60+ (5)

Q3 What is your current clinical role?

Foundation Year (FY 1-2) (1)

Senior house officer (ST 1-2) (2)

Registrar / Specialist registrar (ST 3+) (3)

Consultant / Attending physician / GP (4)

Q4 What medical specialty are you currently working in?

Internal medicine / General hospitalist medicine (1)

Surgery (2)

A&E / Emergency medicine (3)

General practice (GP) / Family medicine (4)

Other, please specify: (5) ________________________________________________

Q5 What type of healthcare facility are you currently practicing in?

Academic / teaching hospital (1)

District general hospital / community hospital (2)

GP practice (3)

Other, please specify: (4) ________________________________________________

Display This Question:

If Q4 = General practice (GP) / Family medicine

Q42 How many clinic sessions do you typically have per week?

None (4)

1-2 (1)

3-4 (2)

5+ (3)

Q6 Where is the hospital/clinic that you are currently working in located?

▼ East of England (1) ... South West (7)

Q8 To the best of your knowledge, how many other healthcare facilities (e.g., clinics, hospitals) does your hospital/clinic typically interact or share patients with?

None (1)

1-2 (2)

3-4 (3)

5+ (4)

I don't know (5)

End of Block: Part 1: Basic Participant Information

Start of Block: Part 2: Clinician EHR User Experience

Q23 Electronic health records (EHRs) are computer systems healthcare providers use to manage a patient’s encounters with the healthcare system. These records can also provide a comprehensive digital view of a patient's clinical history.

Q7 How long have you been using EHRs in your routine clinical work?

Less than 1 year (1)

1-2 years (2)

3-5 years (3)

6-10 years (4)

More than 10 years (5)

Q10 How would you rate your own proficiency using EHR systems?

Beginner (1)

Moderate (2)

Advanced (3)

Expert (4)

Q39 Have you ever received formal training on how to use EHR systems (e.g., courses during medical school, taught by local expert) prior to using them at your workplace?

Yes, please specify: (1) ________________________________________________

No (2)

I don't know (4)

Q11 How often do you personally use EHRs during your routine clinical work?

Less than once a month (1)

At least once a month (2)

At least once a week (3)

Everyday (4)

Q12 Please select the option(s) most appropriate.

 What healthcare tasks can you complete using the EHR at your workplace, and do you use your EHR to perform these tasks often?

|  | I can complete this function with my EHR (1) | I often use my EHR to complete this function (2) |
| --- | --- | --- |
| Hospital administrative processes and reporting (1) |  |  |
| Input orders for investigations/medications (2) |  |  |
| Public health surveillance and reporting (3) |  |  |
| Planning patient disposition/discharges (4) |  |  |
| Aid in clinical decision-making (5) |  |  |
| Retrieve patient’s previous health information (6) |  |  |
| Communicate with other healthcare professionals (7) |  |  |
| Communicate with and support patients (8) |  |  |

Q13 Please select the option most appropriate.
The ability to share clinical information with other healthcare providers is an ability commonly found in EHR systems. In your experience, which of the following clinical information sharing functions are present in the EHR system at your workplace?

|  | Yes (1) | No (2) | I don't know (3) |
| --- | --- | --- | --- |
| I can SEE clinical information inputted by other healthcare providers WITHIN my healthcare setting. (1) |  |  |  |
| I can SEE clinical information inputted by healthcare providers FROM EXTERNAL hospitals/clinics. (2) |  |  |  |
| I can both SEE & EDIT clinical information inputted by other healthcare providers WITHIN my healthcare setting. (4) |  |  |  |
| I can both SEE & EDIT clinical information inputted by healthcare providers FROM EXTERNAL hospitals/clinics. (5) |  |  |  |
| Healthcare providers FROM EXTERNAL hospitals/clinics, can SEE all the clinical information I have inputted. (3) |  |  |  |
| Healthcare providers FROM EXTERNAL hospitals/clinics, can both SEE & EDIT the clinical information I have inputted. (6) |  |  |  |

End of Block: Part 2: Clinician EHR User Experience

Start of Block: Part 3: Implications of EHR Interoperability on Patient Safety & Clinical Care

Q24 The ability to share, retrieve, and interact with clinical information between different healthcare providers and across various healthcare facilities, is a feature found many EHR systems currently in use today. This is often referred to as 'interoperability'.

Q14 In your opinion, how would you describe your overall experience with EHRs and interoperability at your current workplace?

Very Good (1)

Good (2)

Neutral (3)

Bad (4)

Very Bad (5)

Q15 Please select the option most appropriate. 
Have you experienced any of the following interoperability problems when using EHRs at your workplace?:

|  | Always (1) | Most of the time (2) | About half the time (3) | Sometimes (4) | Never (5) |
| --- | --- | --- | --- | --- | --- |
| Difficulty accessing and retrieving clinical information through the EHR systems currently in use. (1) |  |  |  |  |  |
| Difficulty with accessing and retrieving this clinical information negatively affects my day-to-day clinical workflow. (2) |  |  |  |  |  |
| Difficulty with accessing and retrieving this clinical information poses a potential risk to patient safety during my routine shifts in the hospital/clinic. (3) |  |  |  |  |  |
| Difficulty with accessing and retrieving this clinical information negatively impacts my ability to share clinical information with other healthcare professionals. (4) |  |  |  |  |  |
| Difficulty with accessing and retrieving this clinical information negatively impacts my ability to share clinical information with my patients and/or their caregivers. (5) |  |  |  |  |  |
| Thinking about your patients’ expectations regarding accessibility of their health records, do you feel that the EHR systems you currently use allow you to meet these expectations? (6) |  |  |  |  |  |

Q16 When using EHRs, what interoperability difficulties have you encountered which would negatively impact your routine clinical duties or ability to provide care? Please list them here.

________________________________________________________________

________________________________________________________________

________________________________________________________________

________________________________________________________________

________________________________________________________________

Display This Question:

If If When using EHRs, what interoperability difficulties have you encountered which would negatively i... Text Response Is Not Empty

Q17 Thinking about the points you listed above, do you think these present a risk to patient safety?

Yes (1)

No (2)

Maybe (3)

Display This Question:

If Q17 = Yes

Q18 In order of importance, what are the three greatest risks posed to patient safety resultant from issues with EHR interoperability in your clinical workplace?

________________________________________________________________

Q19 Do you regularly encounter any of the following interoperability issues when using EHRs? (Please select all that apply)

- Do not know if the information is available in the EHR system or one that is connected to it. (1)
- Difficulty accessing patient information even when you know that information is available within the system (2)
- Difficulty retrieving patient information you know is available in another healthcare facility frequented by the patient (3)
- Difficulty following up on an order (e.g. test results) you inputted previously (4)
- Difficulty conveying clinical information for another healthcare professional (5)
- Other, please specify: (6) ________________________________________________

Display This Question:

If If Do you regularly encounter any of the following interoperability issues when using EHRs? (Please... q://QID19/SelectedChoicesCount Is Not Empty

And Q4 != General practice (GP) / Family medicine

Q20 When is the lack of interoperable EHRs most impeding to your clinical work during a routine clinical shift? (Please select all options that apply)

- Admitting a new patient from the community (1)
- Receiving a patient from another secondary/tertiary healthcare facility (2)
- During handover of patients from other members within my immediate clinical team (3)
- During handover of a patient from another clinical team/clinician within my hospital (4)
- Following up on an order from another clinical team/clinician in my hospital (5)
- Discharging patient from my hospital back into the community (6)
- Transferring a patient from my hospital to another secondary/tertiary healthcare facility (7)
- Medication reconciliation (9)
- Other, please specify: (8) ________________________________________________

Display This Question:

If Q4 = General practice (GP) / Family medicine

And And Do you regularly encounter any of the following interoperability issues when using EHRs? (Please... q://QID19/SelectedChoicesCount Is Not Empty

Q39 In your experience, which of the following clinical tasks are most commonly impacted by the lack of interoperable EHRs during a routine consultation? (Please select all options that apply)

- Referring patients into hospital (e.g., knowing when the patient is scheduled, ensuring hospital team has up to date GP records) (1)
- Referring patients to allied health services (6)
- Referring patients to another GP clinic (7)
- Receiving outcome of clinical assessments performed in hospital (i.e., clinic letters) (2)
- Receiving results of investigations performed in hospital (3)
- Medication reconciliation (4)
- Other, please specify: (5) ________________________________________________

Q21 In your opinion, what are the top three factors contributing to the current state of EHR interoperability present at your workplace?

________________________________________________________________

Q41 With the onset of the COVID-19 pandemic, did you experience any change (e.g., worsening, improvement) in EHR interoperability at your workplace?

Much better (9)

Somewhat better (10)

About the same (11)

Somewhat worse (12)

Much worse (13)

Q43
Have you experienced any new EHR interoperability issues during the COVID-19 pandemic so far?

Yes, please describe: (1) ________________________________________________

No (2)

I don't know (3)

End of Block: Part 3: Implications of EHR Interoperability on Patient Safety & Clinical Care

Start of Block: Part 4: Costs of Healthcare Resources Used Due to Poor EHR Interoperability

Q35 Issues with EHR interoperability can potentially impact clinicians' ability to provide care, the use of healthcare resources, and patients' length of stay in hospital.

Q28 In your experience, have problems with sharing or retrieving clinical information via EHRs resulted in you having to perform redundant orders/clinical investigations?

Yes (1)

No (2)

I don't know (3)

Q27 What were some of the clinical information sharing problems you have encountered regularly which necessitated repeat orders/investigations?

________________________________________________________________

Q29 On average, how often do you find yourself needing to repeat orders/investigations due to problems with EHR interoperability?

Less than once per week (4)

More than once per week, but less than once per shift/day (5)

1-3 times per day/shift (1)

4-6 times per day/shift (2)

7+ times per day/shift (3)

Q31 Needing to repeat and follow up these orders/investigations frequently hamper me from performing other clinical tasks.

Completely Agree (1)

Somewhat Agree (2)

Not Agree nor Disagree (3)

Somewhat Disagree (4)

Completely Disagree (5)

Q32 On average, how much time per day does repeating and following up on these orders/investigations typically take away from you from completing other clinical duties?

No extra time needed (1)

Less than 5 minutes (2)

5-15 minutes (3)

15-30 minutes (4)

30-60 minutes (5)

1-2 hours (6)

Q33 Please select all relevant options.
What investigations do you find yourself needing to repeat the most often due to EHR interoperability problems (e.g., can't find the records, records are incomplete), and how frequently do you have to do so?

|  | Daily (1) | 4-6 times a week (2) | 2-3 times a week (3) | Once a week (4) | Never (5) |
| --- | --- | --- | --- | --- | --- |
| Baseline panels (i.e., FBC, U&E) (1) |  |  |  |  |  |
| Blood gas (i.e., ABG, VBG) (2) |  |  |  |  |  |
| Cardiac profile (i.e., troponins) (3) |  |  |  |  |  |
| Thyroid panel (i.e., T3/4, TFT) (4) |  |  |  |  |  |
| Liver panel (i.e., LFT) (5) |  |  |  |  |  |
| Lipid profile (i.e., LDL/HDL) (6) |  |  |  |  |  |
| Inflammatory markers (e.g., ESR/CRP) (7) |  |  |  |  |  |
| Blood culture (8) |  |  |  |  |  |
| Urine dipstick (9) |  |  |  |  |  |
| Urinalysis (10) |  |  |  |  |  |
| Urine culture (11) |  |  |  |  |  |
| βHCG pregnancy test (12) |  |  |  |  |  |
| Point of care tests (e.g., Group A Strep, RSV) (13) |  |  |  |  |  |
| Ultrasound (14) |  |  |  |  |  |
| Plain film / X-rays (15) |  |  |  |  |  |
| CT scans (16) |  |  |  |  |  |
| MRI (17) |  |  |  |  |  |
| Other investigations, please specify: (18) |  |  |  |  |  |
| Not Applicable (19) |  |  |  |  |  |

Display This Question:

If Q4 != General practice (GP) / Family medicine

Q34 In your experience, have problems with sharing or retrieving clinical information via EHRs ever necessitated you to keep patients longer in hospital?

Yes (1)

No (2)

I don't know (3)

Q36 In your experience, have problems with sharing or retrieving clinical information via EHRs ever necessitated you to spend more time preparing for clinic?

Yes (1)

No (2)

I don't know (3)

Q44 In your experience, have problems with sharing or retrieving clinical information via EHRs ever necessitated you to prolong the consultation sessions with patients?

Yes (1)

No (2)

I don't know (3)

Display This Question:

If Q34 = Yes

And Q4 = Internal medicine / General hospitalist medicine

Or Q4 = Surgery

Or Q4 = A&E / Emergency medicine

Or Or What medical specialty are you currently working in? Other, please specify: Is Not Empty

Or Q4 = Other, please specify:

Q37 On average, how much delay / prolonged patient stay in hospital are due to problems related to poor EHR interoperability?

No delays caused (1)

Several hours delay (2)

1 night additional stay in hospital (3)

2-3 nights additional stay in hospital (4)

4-6 nights of additional stay in hospital (5)

A week or more stay in hospital (6)

Display This Question:

If Q36 = Yes

Q39 How much extra time do you find yourself spending PREPARING for a routine session/day of clinic due to issues related to EHR interoperability (e.g., cannot find test results, medical history)?

No extra time needed (1)

Less than 5 minutes (4)

5-15 minutes (5)

15-30 minutes (6)

30-60 minutes (7)

More than an hour (8)

Display This Question:

If Q36 = Yes

Q38 How much extra time do you find yourself spending DURING a routine session/day of clinic due to issues related to EHR interoperability?

No extra time needed (1)

Less than 5 minutes (2)

5-15 minutes (3)

15-30 minutes (4)

30-60 minutes (5)

More than an hour (6)

End of Block: Part 4: Costs of Healthcare Resources Used Due to Poor EHR Interoperability

### Appendix 2

Table 12: Aggregated investigation categories

| **Investigation categories** | **Included investigations** |
| --- | --- |
| Bloodwork | Baseline panels (*e.g.,* FBC, U&E)  Cardiac profile (*e.g.,* troponin)  Thyroid panel (*e.g.,* TFT)  Liver panel (*e.g.,* LFT)  Lipid profile (*e.g.,* LDL/HDL)  Inflammatory markers (*e.g.,* ESR/CRP) |
| Blood gases | ABG/VBG |
| Urine-based | Urine dipstick  Urinalysis |
| Infectious panels | Urine culture  Blood culture |
| Radiology | Ultrasound  Plain films/X-rays  CT scans  MRI |
| Miscellaneous investigations | Point-of-care tests (*e.g.,* Group A Strep, RSV)  βHCG pregnancy test  Other investigations (please specify) |
| Non-applicable | Non-applicable |

### Appendix 3

Table 10: Participant characteristics (Gender) at the beginning and end of the survey. The chi-square statistic is 3.3191. The p-value is .19022. The result is not significant at p < 0.05.

| **Gender** | **Number of responses, at the beginning of survey, n (%)** | **Number of responses, at the end of survey, n (%)** | **Number of missing responses by the end of the survey, n (%)** |
| --- | --- | --- | --- |
| Male | 313 (51.91%) | 124 (56.88%) | 189 (49.22%) |
| Female | 282 (46.77%) | 92 (42.20%) | 190 (49.48%) |
| Missing | 8 (1.33%) | 2 (0.92%) | 5 (1.30%) |

Table 11: Participant characteristics (Age band) at the beginning and end of the survey. The chi-square statistic is 19.2532. The p-value is .000701. The result is significant at p < .05.

| **Age band (years)** | **Number of responses, at the beginning of survey, n (%)** | **Number of responses, at the end of survey, n (%)** | **Number of missing responses by the end of the survey, n (%)** |
| --- | --- | --- | --- |
| Under 30 | 122 (20.23%) | 31 (14.22%) | 91 (75.60%) |
| 30-39 | 224 (37.15%) | 71 (32.57%) | 153 (12.70%) |
| 40-49 | 145 (24.05%) | 60 (27.52%) | 85 (7.05%) |
| 50-59 | 84 (13.93%) | 43 (19.72%) | 41 (3.40%) |
| 60+ | 28 (4.64%) | 13 (5.96%) | 15 (1.24%) |

Table 12: Participant characteristics (Clinical role) at the beginning and end of the survey. The chi-square statistic is 26.258. The p-value is < 0.00001. The result is significant at p < .05.

| **Clinical role** | **Number of responses, at the beginning of survey, n (%)** | **Number of responses, at the end of survey, n (%)** | **Number of missing responses by the end of the survey, n (%)** |
| --- | --- | --- | --- |
| Foundation year (FY1-2) | 59 (9.78%) | 8 (3.67%) | 51 (13.35%) |
| Senior house officers (ST1-2) | 98 (16.25%) | 26 (11.93%) | 72 (18.70%) |
| Registrar/Senior registrar (ST3+) | 180 (29.85%) | 64 (29.36%) | 116 (30.13%) |
| Consultant/GP | 266 (44.11%) | 120 (55.05%) | 146 (37.92%) |

Table 13: Participant characteristics (Location of practice) at the beginning and end of the survey. The chi-square statistic is 3.016. The p-value is < 0.806818. The result is not significant at p < .05.

| **Location of practice** | **Number of responses, at the beginning of survey, n (%)** | **Number of responses, at the end of survey, n (%)** | **Number of missing responses by the end of the survey, n (%)** |
| --- | --- | --- | --- |
| East of England | 52 (9.39%) | 16 (7.62%) | 36 (11.25%) |
| London | 155 (27.98%) | 64 (30.48%) | 91 (28.44%) |
| Midlands | 54 (9.75%) | 19 (9.05%) | 35 (10.94%) |
| North East and Yorkshire | 91 (16.43%) | 37 (17.62%) | 54 (16.88%) |
| North West | 131 (23.65%) | 46 (21.90%) | 85 (26.56%) |
| South East | 30 (5.42%) | 11 (5.24%) | 19 (5.94%) |
| South West | 41 (7.40%) | 17 (8.10%) | 24 (7.50%) |

Table 14: Participant characteristics (Medical specialty training) at the beginning and end of the survey. The chi-square statistic is 47.304. The p-value is .0001. The result is significant at p < .05.

| **Medical specialty training** | **Number of responses, at the beginning of survey, n (%)** | **Number of responses, at the end of survey, n (%)** | **Number of missing responses by the end of the survey, n (%)** |
| --- | --- | --- | --- |
| Internal medicine and medicine subspecialties | 226 (37.67%) | 91 (42.13%) | 135 (35.16%) |
| Surgery and surgical subspecialties | 123 (20.50%) | 66 (30.56%) | 57 (14.84%) |
| A&E | 56 (9.33%) | 5 (2.31%) | 51 (13.28%) |
| Anaesthesia | 48 (8%) | 8 (3.70%) | 40 (10.42%) |
| Paediatrics | 43 (7.17%) | 17 (7.87%) | 26 (6.77%) |
| Psychiatry | 41 (6.83%) | 11 (5.09%) | 30 (7.81%) |
| GP/Family medicine | 50 (8.33%) | 14 (6.48%) | 36 (9.38%) |
| Other | 13 (2.17%) | 4 (1.85%) | 9 (2.34%) |

Table 15: Participant characteristics (Healthcare facility type) at the beginning and end of the survey. The chi-square statistic is 2.1353. The p-value is .544808. The result is not significant at p < .05.

| **Type of healthcare facility** | **Number of responses, at the beginning of survey, n (%)** | **Number of responses, at the end of survey, n (%)** | **Number of missing responses by the end of the survey, n (%)** |
| --- | --- | --- | --- |
| Academic/teaching hospital | 290 (48.09%) | 112 (51.38%) | 178 (46.23%) |
| District general/community hospital | 244 (40.46%) | 84 (38.53%) | 160 (41.56%) |
| GP practice | 42 (6.97%) | 12 (5.50%) | 30 (7.79%) |
| Other | 27 (4.48%) | 10 (4.59%) | 17 (4.42%) |

Table 16: Participant characteristics (Number of organisations interacting with) at the beginning and end of the survey. The chi-square statistic is 4.8779. The p-value is .180963. The result is not significant at p < .05.

| **Number of organisations interacting with** | **Number of responses, at the beginning of survey, n (%)** | **Number of responses, at the end of survey, n (%)** | **Number of missing responses by the end of the survey, n (%)** |
| --- | --- | --- | --- |
| None | 7 (1.45%) | 2 (1.06%) | 5 (1.69%) |
| 1-2 | 58 (11.98%) | 17 (8.99%) | 41 (13.90%) |
| 3-4 | 144 (29.75%) | 52 (27.51%) | 92 (31.19%) |
| 5+ | 275 (56.82%) | 118 (62.43%) | 157 (53.22%) |

Table 17: Participant characteristics (Received formal EHR training) at the beginning and end of the survey. The chi-square statistic is 36.4961. The p-value is < .00001. The result is significant at p < .05.

| **Received formal EHR training** | **Number of responses, at the beginning of survey, n (%)** | **Number of responses, at the end of survey, n (%)** | **Number of missing responses by the end of the survey, n (%)** |
| --- | --- | --- | --- |
| Yes | 256 (56.51%) | 88 (41.51%) | 168 (69.71%) |
| No | 197 (43.49%) | 124 (58.49%) | 73 (30.29%) |

Table 18: Participant characteristics (Self-reported EHR proficiency level) at the beginning and end of the survey. The chi-square statistic is 3.0306. The p-value is .386936. The result is not significant at p < .05.

| **Self-reported EHR proficiency level** | **Number of responses, at the beginning of survey, n (%)** | **Number of responses, at the end of survey, n (%)** | **Number of missing responses by the end of the survey, n (%)** |
| --- | --- | --- | --- |
| Beginner | 23 (4.96%) | 14 (6.42%) | 9 (3.66%) |
| Moderate | 235 (50.65%) | 109 (50%) | 126 (51.22%) |
| Advanced | 167 (35.99%) | 74 (33.94%) | 93 (37.80%) |
| Expert | 39 (8.41%) | 21 (9.63%) | 18 (7.32%) |

Table 19: Participant characteristics (Frequency of EHR use) at the beginning and end of the survey. The chi-square statistic is 0.4191. The p-value is .936259. The result is not significant at p < .05.

| **Frequency of EHR use** | **Number of responses, at the beginning of survey, n (%)** | **Number of responses, at the end of survey, n (%)** | **Number of missing responses by the end of the survey, n (%)** |
| --- | --- | --- | --- |
| Less than once a month | 2 (0.43%) | 1 (0.46%) | 1 (0.41%) |
| At least once a month | 7 (1.51%) | 1 (0.46%) | 1 (0.41%) |
| At least once a week | 31 (6.68%) | 13 (5.96%) | 18 (7.47%) |
| Every day | 424 (91.38%) | 203 (93.12%) | 221 (91.70%) |

### Appendix 4

Table 20: Participant responses to whether limited EHR interoperability resulted in redundant investigations, prolonged patient length of stay in hospital, and more time needed for preparing + conducting consultations.

|  | **n** | **Of those who responded, (%)** | **Out of total number of participants, (%)** |
| --- | --- | --- | --- |
| Have problems sharing or retrieving clinical information via EHRs resulted in you having to perform redundant orders/clinical investigations? |  |  |  |
| Yes | 217 | 73.06% | 34.12% |
| No | 80 | 26.94% | 12.58% |
| Missing | 339 | N/A | 53.30% |
| Have problems sharing or retrieving information via EHRs necessitated keeping patients in hospital longer? |  |  |  |
| Yes | 142 | 64.25% | 22.33% |
| No | 79 | 35.75% | 12.42% |
| Missing | 415 | N/A | 65.25% |
| Have problems sharing or retrieving information via EHRs necessitated you spending more time preparing for clinic consultations? |  |  |  |
| Yes | 224 | 81.16% | 35.22% |
| No | 52 | 18.84% | 8.18% |
| Missing | 360 | N/A | 56.60% |
| Have problems sharing or retrieving information via EHRs necessitated you prolonging the actual consultation session itself? |  |  |  |
| Yes | 47 | 15.72% | 7.39% |
| No | 252 | 84.28% | 39.62% |
| Missing | 337 | N/A | 52.99% |

### Appendix 5

Table 21: Cross tabulation of perceptions of current state of EHR interoperability by age group

|  | **Very Bad, n (%)** | **Bad, n (%)** | **Neutral, n (%)** | **Good, n (%)** | **Very Good, n (%)** |
| --- | --- | --- | --- | --- | --- |
| Under 30 | 10 (13.33%) | 13 (17.33%) | 22 (29.33%) | 22 (29.33%) | 8 (10.67%) |
| 30-39 | 17 (12.06%) | 30 (21.28%) | 34 (24.11%) | 52 (36.88%) | 8 (5.67%) |
| 40-49 | 14 (12.50%) | 29 (25.89%) | 33 (29.46%) | 28 (25.00%) | 8 (7.14%) |
| 50-59 | 10 (15.38%) | 12 (18.46%) | 23 (35.38%) | 17 (26.15%) | 3 (4.62%) |
| 60+ | 4 (19.05%) | 6 (28.57%) | 5 (23.81%) | 5 (23.81%) | 1 (4.76%) |

### Appendix 6

Table 22: Repeat diagnostic investigations performed because of poor EHR interoperability (non-aggregated figures) from total number of responses.

|  | **Daily**  **n (%)** | **4-6 times a week n (%)** | **2-3 times a week n (%)** | **Once a week n (%)** | **Never n (%)** |
| --- | --- | --- | --- | --- | --- |
| Baseline panels (*i.e.,* FBC) | 12 (4.29%) | 12 (4.29%) | 32 (11.43%) | 97 (34.64%) | 127 (45.36%) |
| Cardiac profile (*i.e.,* troponins) | 1 (0.38%) | 1 (0.38%) | 10 (1.57%) | 37 (13.96%) | 216 (81.51%) |
| Thyroid panel (*i.e.,* TFT) | 4 (1.48%) | 2 (0.74%) | 9 (3.33%) | 68 (25.19%) | 187 (69.26%) |
| Liver panel (*i.e.,* LFT) | 4 (1.49%) | 1 (0.37%) | 15 (5.58%) | 80 (29.74%) | 169 (62.83%) |
| Lipid profile (*i.e.,* LDL/HDL) | 4 (3.37%) | 1 (0.37%) | 9 (3.37%) | 48 (17.98%) | 205 (76.78%) |
| Inflammatory markers (*i.e.,* ESR/CRP) | 8 (2.97%) | 6 (2.23%) | 13 (4.83%) | 84 (31.23%) | 158 (58.74%) |
| Blood gas (i.e., ABG, VBG) | 3 (1.10%) | 3 (1.10%) | 11 (4.04%) | 48 (17.65%) | 207 (76.10%) |
| Urine dipstick | 12 (4.62%) | 11 (4.23%) | 40 (15.38%) | 61 (23.46%) | 136 (52.31%) |
| Urinalysis | 11 (4.21%) | 9 (3.45%) | 25 (9.58%) | 68 (26.05%) | 148 (56.70%) |
| Blood culture | 2 (0.76%) | 0 (0.00%) | 5 (1.91%) | 64 (24.43%) | 191 (72.90%) |
| Urine culture | 5 (1.92%) | 5 (1.92%) | 16 (6.13%) | 62 (23.75%) | 173 (66.28%) |
| Ultrasound | 6 (2.21%) | 3 (1.10%) | 18 (6.62%) | 75 (27.57%) | 170 (62.50%) |
| Plain film/X-rays | 5 (1.85%) | 4 (1.48%) | 13 (4.81%) | 93 (34.44%) | 155 (57.41%) |
| CT scans | 4 (1.46%) | 4 (1.46%) | 12 (4.38%) | 82 (29.93%) | 172 (62.77%) |
| MRI | 4 (1.50%) | 3 (1.12%) | 7 (2.62%) | 66 (24.72%) | 187 (70.04%) |
| Point-of-care tests | 4 (1.57%) | 4 (1.57%) | 7 (2.75%) | 32 (12.55%) | 208 (81.57%) |
| βHCG pregnancy test | 8 (3.07%) | 3 (1.15%) | 13 (4.98%) | 38 (14.56%) | 199 (76.25%) |
| Other investigations | 5 (4.07%) | 2 (1.63%) | 4 (3.25%) | 24 (19.51%) | 88 (71.54%) |
| Not applicable | 13 (11.50%) | 1 (0.88%) | 2 (1.77%) | 5 (4.42%) | 92 (81.42%) |

### Appendix 7

*Table 23 Breakdown of doctors reporting negative experiences of EHR interoperability at their workplace, with crude and adjusted odds ratios (n=414). *** p≤ 0.01, **p ≤ 0.05, *p ≤ 0.1.*

| Characteristics | Odds ratio (95% CI) | P-value | Adjusted odds ratio (95% CI) | P-value |
| --- | --- | --- | --- | --- |
| **Age Group** |  |  |  |  |
| <29 | Reference | N/A | Reference | N/A |
| 30-39 | 0.885 (0.484 - 1.617) | 0.717 | 0.566 (0.216 - 1.482) | 0.246 |
| 40-49 | 0.710 (0.381 - 1.321) | 0.271 | 0.307* (0.093 - 1.013) | 0.052 |
| 50-59 | 0.865 (0.425 - 1.759) | 0.257 | 0.381 (0.106 - 1.376) | 0.142 |
| 60+ | 0.487 (0.181 - 1.306) | 0.342 | 0.155 (0.033 - 0.735) | 0.019 |
| **Gender** |  |  |  |  |
| Female | Reference | N/A | Reference | N/A |
| Male | 0.668* (0.442 - 1.009) | 0.055 | 0.698 (0.444 - 1.099) | 0.120 |
| **Level of Clinical Training** |  |  |  |  |
| Consultant | Reference | N/A | Reference | N/A |
| Registrar / Specialist Registrar (ST 3+) | 0.942 (0.588 - 1.509) | 0.805 | 0.514 (0.225 - 1.174) | 0.114 |
| Senior House Officer (ST 1-2) | 1.362 (0.724 - 2.563) | 0.338 | 0.497 (0.159 - 1.554) | 0.230 |
| Foundation Year (FY 1-2) | 0.731 (0.346 - 1.545) | 0.412 | 0.246** (0.061 - 1.000) | 0.050 |
| **Specialty Training** |  |  |  |  |
| Internal medicine / Medicine subspecialties | Reference | N/A | Reference | N/A |
| Surgery / Surgical subspecialties | 0.613* (0.363 - 1.036) | 0.068 | 0.638 (0.365 - 1.116) | 0.115 |
| A&E / Emergency medicine | 0.867 (0.416 - 1.809) | 0.704 | 0.735 (0.340 - 1.588) | 0.434 |
| Anaesthesia | 1.735 (0.704 - 4.277) | 0.232 | 1.974 (0.768 - 5.074) | 0.158 |
| Paediatrics | 0.486* (0.210 - 1.123) | 0.091 | 0.365** (0.147 - 0.904) | 0.029 |
| Psychiatry | 0.648 (0.256 - 1.636) | 0.358 | 0.589 (0.218 - 1.589) | 0.296 |
| General practice/Family Medicine | 3.011** (1.106 - 8.203) | 0.031 | 2.729* (0.950 - 7.841) | 0.062 |
| Other | 0.648 (0.140 - 3.002) | 0.579 | 0.830 (0.147 - 4.700) | 0.833 |
| **Years of Using EHR** |  |  |  |  |
| 3-5 years | Reference | N/A | Reference | N/A |
| Less than 1 year | 1.219 (0.445 - 3.338) | 0.700 | 2.295 (0.693 - 7.606) | 0.174 |
| 1-2 years | 0.688 (0.389 - 1.219) | 0.200 | 0.693 (0.356 - 1.352 | 0.282 |
| 6-10 years | 0.640 (0.366 - 1.120) | 0.118 | 0.685 (0.374 - 1.257) | 0.222 |
| More than 10 years | 0.797 (0.435 - 1.461) | 0.463 | 0.773 (0.373 - 1.602) | 0.488 |
| **Self-reported EHR Proficiency** |  |  |  |  |
| Moderate | Reference | N/A | Reference | N/A |
| Beginner | 0.489 (0.202 - 1.185) | 0.113 | 0.419* (0.156 - 1.124) | 0.084 |
| Advanced | 0.968 (0.618 - 1.518) | 0.889 | 0.962 (0.581 - 1.590) | 0.879 |
| Expert | 0.604 (0.299 - 1.220) | 0.160 | 0.586 (0.264 - 1.303) | 0.190 |
